# Compliance with the provisions related to higher educational institutes of anti-tobacco/smoking law by institutes of national importance in India

**DOI:** 10.1101/2023.02.13.23285898

**Authors:** Raja Singh

**Affiliations:** Visiting Faculty, Department of Architecture, School of Planning and Architecture, New Delhi; Built Environment and Public Health Research Fellow, Tathatara Foundation, India; Advisor, ISAC Centre for Built Environment Policy, India

**Keywords:** Anti-smoking law, Anti-tobacco law, educational institutions, signboards, Institutes of National Importance, India, Right to Information Act

## Abstract

India formulated an anti-tobacco and anti-smoking law in 2003 in response to its resolutions in the United Nations’ bodies. This law has been detailed subsequently to make it focussed on educational institutions, which are supposed to perform on-ground action. The first is to put up signboards prohibiting the sale of cigarettes and tobacco products within 100 yards, the second is prohibiting smoking within the campus, and the third is implementing the law and collecting fines from the offenders. The focus of this paper is on India’s premier educational institutions called the ‘Institutions of National Importance’ by the Indian legislature. These are India’s public institutions which have a focus of the government in making them high quality. The paper checked for the compliance of the Indian anti-smoking and anti-tobacco laws in 79 of these institutions. The Right to Information Act, 2005, India’s transparency law, was used to file applications for information, and certified information from the institutions was collected and reported. The results show an overall weak compliance with the law. India’s health regulators and educational watchdogs must implement anti-smoking and anti-tobacco laws strictly in Indian educational institutions.

##### What is already known on this topic

There is noncompliance of India’s anti-smoking /anti-tobacco legislation, esp. signboards.

##### What this study adds

This study adds pan India based compliance at higher educational institutions with a study on fine collection and signboard compliance.

##### How this study might affect research, practice or policy

This study is a wakeup call for Indian educational institutions as India has decentralised implementation of the anti-smoking law to educational institutions and they must perform this duty well.

## Introduction

India confers the status of Institute of National Importance to premier public higher education institutes through legislative action by an act of parliament. This institution is defined as serving ‘as a pivotal player in developing highly skilled personnel within the specific region of the country’ [1,2]. These institutes receive special recognition, higher autonomy and funding from the Government of India. They are equivalent to universities in themselves and are not affiliated with any other mother university but are autonomously governed by a common council to oversee and standardise these. There are, as of 2023, 161 Institutes of National importance in India, and these include the Indian Institutes of Technologies and National Institutes of Technology and the All India Institutes of Medical Sciences, among other important institutes [1]. These are the crème-de-la-crème of educational institutes in India, and it is supposed that actions in these reverberate across the country. With this respect, their compliance with the law will also serve as a barometer of compliance by educational institutes in India.

India is governed as a democratic country with the rule of law. The parliament legislates and creates statutes which are called Acts. Many of these laws are created in response to the global commitments that the country makes on the level of International diplomacy and politics. Some examples are the Right for Persons with Disabilities Act 2016, which was made after a resolution of the United Nations in which India was a signatory. In the same respect, India has also enacted the Cigarettes and Other Tobacco Products (Prohibition of Advertisement and Regulation of Trade and Commerce, Production, Supply and Distribution) Act, 2003. This was done in response to the resolution in the 39^th^ and the 43^rd^ World Health Assembly resolutions [3]. The 43^rd^ World Assembly resolution was all the more important as it focussed on protecting children, specifically among others. The Cigarettes and Other Tobacco Products (Prohibition of Advertisement and Regulation of Trade and Commerce, Production, Supply and Distribution) Act, 2003 or the COTPA Act, 2003 [4]hereinafter was created with the preamble to create a law which is ‘ a comprehensive law on tobacco in the public interest and to protect the public health and ‘to prohibit the consumption of cigarettes and other tobacco products which are injurious to health with a view of achieving the public health as enjoined by Article 47’ of the Indian Constitution.

The first landmark of the COTPA Act 2003 is that it out-rightly states in Section 4 that

> *‘No person shall smoke in any public place.’*

This is made to include all public places with certain exceptions where designated smoking zones can be created.

In India, the Acts are general are the general substantive principles which are further procedurally detailed out through rules and notifications through the official gazette of India. The COTPA Act 2003 is the substantive element of the anti-smoking law in India, and this has been further detailed out to lay procedure through various rules notified thereafter by the Central Government. The provisions of the scope of these rules are always derived from the main law itself, which is COTPA Act 2003 in this case.

For the young population, as we spoke before, the COTPA Act 2003 specifies in Section 6 as follows:

> *‘No person shall sell, offer for sale, or permit the sale of, cigarette or any other tobacco product-*

a. *To any person who is under eighteen years of age, and*
b. *In an area within a radius of one hundred yards of any educational institution*

In order to procedurally outline the above substantive principle, the Central government notified the Cigarettes and other Tobacco Products (Display of Boards by Educational Institutions) Rules 2009 [5], which clearly instruct, under its Section 3, educational institutes the following:

> *3. Display of Board by Educational Institutions. (1) The owner or manager or any person in-charge of affairs of the educational institution shall display and exhibit a board at a conspicuous place outside the premises, prominently stating that sale of cigarettes and other tobacco products in an area within a radius of one hundred yards of educational institution is strictly prohibited and that it is an offense under Section 24 of the Act with fine which may extent to two hundred rupees.’*

The above has been made an offence under Section 24 of the COTPA Act, 2003.

The government has clearly defined the definition of educational institutes in Section 2(b) of the Prohibition of Sale of Cigarettes and other Tobacco Products Around Educational Institutions Rules, 2004, which is stated as ‘means places/centres where educational instructions are imparted according to the specific norms and include schools, colleges and institutions of higher learning established or recognised by an appropriate authority;’ The distance measurement has been fine-tuned in Section 3(2) of the same as to be measured ‘be measured radially starting from the outer limit of boundary wall, fence or as the case may be, of the educational institution.’

The government has further notified the Prohibition of Smoking in Public Places Rules, 2008, which further strengthens the anti-smoking regime. To decentralise the process of enforcement, the Central government has distributed the power to book and compound the offenders under the COTPA Act, 2003. The list of officers authorised to compound is provided in Schedule III of the Prohibition of Smoking in Public Places Rules, 2008. In the case of an educational institution, the Principal/Teacher /Director/Medical Superintendent/Head of the Institution is the person authorised to take action and collect the fines. This automatically means that a record of the instances of violation of the COTPA Act 2003 and its rules, along with the collection of fines, becomes the responsibility of the head of the institution. If no record is maintained, it is safe to either assume that no instances of smoking have ever taken place or no fines have ever been collected. The worst assumption, which may be true, is if the head of the institution never complied with the provisions of the said law.

The above notification, i.e., the Prohibition of Smoking in Public Places Rules, 2008 [6], also puts the responsibility on the educational institute, which falls under the category of public places, to put boards which prohibit people from smoking. The verbatim provision of the law is given in Section 3(b), which is as follows:

> *The board as specified in Schedule II is displayed prominently as the entrance of the public place, in case there are more than one entrance at each such entrance and conspicuous place 9s) inside. In case there are more than one floor, at each floor including staircase and entrance to the lift/s at each floor*.

This means that the boards have to be placed at the following in a public place, including an educational institution:

1. The entrance of the public place. In case of more than one entrance, at each such entrance.
2. On each floor, including at the staircase or lift entrance of each floor.

It is required that the board be placed at a conspicuous place where it is effortlessly visible to general visitors at first glance.

The Prohibition of Smoking in Public Places Rules, 2008 have also, to remove any confusion or doubt, made the specification of the required board as part of the statute.

Schedule II, with the details of the boards, is as follows:

1. *The board shall be of a minimum size of 60 cm by 30 cm of white background*.
2. *It shall contain a circle of no less than 15 cm outer diameter with a red perimeter of no less than 3 cm wide with a picture, in the centre, of a cigarette or ‘beedi*^*1*^*’ with black smoke and crossed by a red hand*.
3. *The width of the red band across the cigarette shall equal the width of the red perimeter*.
4. *The board shall contain the warning “No Smoking Area-Smoking Here is an Offence” in English or one Indian language, as applicable*.

It is often stated that the COTPA Act 2003 may not have the required teeth as the maximum fine is rupees 200, which is about 2 dollars or a little more. This may not set a deterrent enough. Another route taken by the legislature has been the enactment of the Juvenile Justice (Care and Protection of Children) Act, 2015 [7], which has made selling tobacco products to a child punishable with a rigorous imprisonment of seven years and has also imposed a fine which may extend up to one lakh rupees which are about 1200 USD.

The question of the effectiveness of the prohibition within 100 yards of educational institutes is beyond this study. The scope is largely on whether there is compliance with the various parameters of the COTPA Act 2003 and the two rules notified under it. The major focus is on the presence of the required signboards, which create awareness and serve as a nudge to the users to not smoke or to vendors to not sell cigarettes and other tobacco products to users. Apart from this, the focus is also on whether there is an administrative measure with respect to the enforcement of warnings and fines for users who may be using cigarettes and other tobacco products within educational institutes.

### Aim

To study the compliance status of the anti-smoking/tobacco law in India’s premier higher educational institutions.

### Objectives

1. To study whether there is compliance in terms of the installation of the signboards that prohibit the sale of cigarettes and tobacco within 100 yards of the premier institutions in India.
2. To study the compliance in terms of the installation of signboards prohibiting smoking within educational institutions.
3. To study whether the premier educational institutions are enforcing the prohibition of smoking within the campus by imposing fines on violators of the ban on smoking.
4. To know the approach of the premier educational institutes in India towards the presence of tobacco vendors within 100 yards of the educational institutions.
5. To study whether the premier educational institutions are undertaking any awareness activities to curb smoking and tobacco use.

### Need for the study

The anti-smoking law in India, called the COTPA Act 2003, is a remarkable example of anti-smoking and tobacco legislation. The landmark feature of the rules drafted after their enactment is the decentralisation of the enforcement and implementation that has been done in India. This means that the on-ground work is performed not directly by the government but is distributed to the organisation where the interaction takes place. This means that no special force or authority as such is responsible, but a multitude of on-ground agencies are involved. For example, in the Prohibition of Smoking in Public Places Rules, 2008, the responsibility of the implementation, including the collection of fines, lies with the Head of the Institution of the educational institute. In another example, even the board that is to be placed outside of the educational institute is to be done by the educational institute and not necessarily by the municipality. This means the implementation is outsourced to the end agency.

The placement of required anti-tobacco health messaging boards, collection of fines, and awareness measures performed by educational institutions need to be checked as it reflects the performance of the law.

There have been multiple studies that have looked into this issue and checked for related issues, but the uniqueness of this study lies in the number of educational institutions, and the pan-India geographic approach, which touches most, if not all, the states and union territories in India. The other factor highlighting the importance of this study is that it is performed on Institutes of National Importance, which are the premier publically financed Institutes in India. No such study in India has been performed at this scale across India in such premier institutes. Another unique element is the use of India’s transparency law for obtaining information required for this study.

## Materials and Methods

The following steps were performed to undertake this study:

1. A need for the study was established through literature.
2. A proforma for the information required was drafted with respect to the objectives stated above.
3. Applications under the Right to Information Act 2005 were drafted and sent to the premier educational institutes.
4. The information provided by the institute under the sign and seal through the authorised channel was collected and compiled.
5. Institutes where information was not provided, incomplete information was provided, or any clarification was sought were appealed under the provisions of the Right to Information. The information so derived was compiled along with the information received earlier.
6. The information collected was reported.

The educational institutions that were requested for information are as follows:

**Table 1:** Table stating the number of each type of educational institution that provided information under the Right to Information Act, 2005.

| S. No. | Category of Educational Institution | Number of Institutions of this type | Remarks |
| --- | --- | --- | --- |
| 1. | Indian Institutes of Management | 20 | These are the premier public-funded management institutions in India. |
| 2. | Indian Institutes of Technology | 22 | These are the public-funded premier engineering-based educational institutes in India. |
| 3. | National Institutes of Technology | 30 | These are premier engineering educational institutions in India of public nature. |
| 4. | Indian Institutes of Science Education and Research | 7 | These are STEM-based fundamental science-based educational institutions in India. |
| Total |  | 79 |  |

The information sought from the educational institutions is given in Table 2 below:

**Table 2:** Table with the information requested from institutes of national importance with the rationale, through the Right to Information Act, 2005.

| S. No. | Information Requested | Why was the information requested for |
| --- | --- | --- |
| 1 | Please provide the record of the presence, number and locations of boards installed at a conspicuous place outside the educational institution in compliance with the Prohibition of Sale of Cigarettes and other Tobacco Products around Educational Institutions Rules, 2004, which prohibits the sale of cigarettes and tobacco products in an area within a radius of one hundred yards. (See Section 3(1) of the Prohibition of Sale of Cigarettes and other Tobacco Products around Educational Institution Rules, 2004). | The law provides that all educational institutions in India put boards that prohibit the sale of cigarettes and tobacco within 100 yards. This information checked the compliance of the same as it is specific to the sale of cigarettes and tobacco products. |
| 2 | Please provide the record of the number and location of boards installed at the entrances stating No Smoking Area-Smoking Here is an offence as per Section 3(b) and Schedule II of the Prohibition of Smoking in Public Places Rules, 2008. | Apart from the boards mentioned in point 1, all public places have to have a board stating 'No Smoking Area'. This information is checked for compliance with the same as it prohibits the use and |
|  |  | not the sale. |
| 3 | Please provide the information and records of the total instances of fines collected/offences compounded/offences recorded/warnings issued with respect to Section 5 of the Prohibition of Smoking in Public Places Rules, 2008, which states the authorised officers who are competent to act under the act as per Schedule III of Prohibition of Smoking in Public Places Rules, 2008. The period of information is from 2012 till date.<br>Note: The Principal/Teacher/Director/MS/Head of the Institution is a person authorised to take action under the act. | The collection of fines has been decentralised by law. This means the Head of the Institution (Public/Private) is empowered to enforce no smoking and no tobacco use within educational institutions. This can be done without involving law enforcement agencies. The collection of fines is a metric in this direction. |
| 4 | Please provide the number of vendors of cigarettes and other tobacco products within a distance of 100 yards from the outer boundary of the educational institute. | Since the sale of cigarettes and tobacco products is prohibited within 100 yards, the role the educational institute plays in enforcing this is to be checked by this information request. |
| 5 | Please provide a list of events/initiatives/activities/circulars/anything on record where the educational institution has taken steps to prevent the use of cigarettes and tobacco products | Educational institutes play a role in shaping the minds of students and society. Awareness of tobacco and cigarette smoking plays a key role and is checked by this information request. |

The sample size justification of the study requires first finding out the total number of Institutes of National Importance in the country. The total number of institutes of national importance in the country is 161[1]. The sample for which the study has been done is 79 institutes. This means that the Confidence level is 95% +/- 7.9% margin of error.

The process of collecting information through the channel of the Right to Information Act 2005 required going back and forth with the educational institutions, and in many cases, an appeal had to be made to get the maximum possible information.

### Non-requirement of ethics clearance for this study

This study included no questionnaire and no human subjects. No employee, visitor, or staff was contacted directly for this study. This study used information available in the public domain through the Right to Information Act 2005, where an application was made under an appropriate section of the Act, and the information was supplied. The information provided was signed and certified by the airport through a senior official and released into the public domain. The Right to Information Act 2005 allows for the provision of only such information that is not the third-party or personal information of any individual. This prevents any information about any human subject. The use of information in this study is available under the public domain and has no human subjects and is thus exempt from review under the ‘Indian Council of Medical Research: National Ethics Guidelines for Biomedical and Health Research involving Human Participants’[8]. The study is in the domain of well-established anti-smoking law legislated and notified in India to see whether it is being complied with in letter or spirit. According to the above-mentioned scope, this study does not require Ethics Committee Approval or its equivalent Institutional Review Board Approval. The author declares the same.

## Results

Out of the 79, 31 educational institutions had the requisite board outside of the campus in the format which prohibits the sale of cigarettes and tobacco products in an area within a radius of one hundred yards. (See Section 3(1) of the Prohibition of Sale of Cigarettes and other Tobacco Products around Educational Institution Rules, 2004). 8 out of the 79 had boards placed at the main entry-exit points, but they were not in the format as per mentioned in the statute. 17 confessed to having no such board placed. The remaining 23 educational institutes of national importance simply denied the information regarding the presence of the board, which prohibits the sale of cigarettes and tobacco within 100 yards of educational institutes. See Table 3.

**Table 3:** Table showing the summary of the information provided by the Placement of Boards at a conspicuous place outside the educational institution in compliance to the Prohibition of Sale of Cigarettes and other Tobacco Products around Educational Institutions Rules, 2004, which prohibits the sale of cigarettes and tobacco.

| No. of Institutions that had placed the board in the requisite format | No. of institutions that had placed boards but were not in the format required | No. of institutions that had not put a board | No. of institutions that did not provide the information. | Total number of institutions |
| --- | --- | --- | --- | --- |
| 31 [39.2%] | 8 [10.1%] | 17 [21.5%] | 23 [29.1%] | 79 |

With respect to the provision of a ‘No Smoking Area-Smoking Here is an Offence’ board in the same or similar format, 58 educational institutes had the requisite boards in the campus. 2 institutions stated that they had initiated the process of installations of the boards, which was after the application for information was filed by the author. 3 educational institutions confessed to having no boards installed. 16 educational institutions denied the information by not providing a reply. See Table 4.

**Table 4:** The record of the number and location of boards installed by educational institutions stating No Smoking Area-Smoking Here is an offence as per Section 3(b) and Schedule II of the Prohibition of Smoking in Public Places Rules, 2008.

| No. of Institutions that had placed the No Smoking boards in the requisite format | No. of institutions that stated that they had initiated the process of installation of the boards. | No. of institutions that had not put a board | No. of institutions that did not provide the information. | Total number of institutions |
| --- | --- | --- | --- | --- |
| 58 [73.4%] | 2 [2.5%] | 3 [3.8%] | 16 [20.25%] | 79 |

With respect to the educational institutions penalising people smoking in campus by issuing warnings, collecting fines and reporting offenders, the educational institutions had the following response. 31 educational institutions reported no incidence of smoking and therefore collected no fines. This, first of all, means that they had a mechanism to record such activities and had compliance in the first place. 14 out of the 79 institutions had taken cognisance and had either issued warnings of the incidents of smoking/tobacco use within the campus, and most out of the 14 had collected fines, as required by the law. 34 out of the 79 educational institutions denied the information by either not providing a requisite reply or by stating that this information was not part of their records. See Table 5.

**Table 5:** Summary of the number of institutions with the information and records of the total instances of fines collected/offences compounded/offences recorded/warning issued in respect to Section 5 of the Prohibition of Smoking in Public Places Rules, 2008.

| No. of Institutions that reported no instance of smoking and/or fine imposed | No. of institutions that took cognisance and either issued warnings or collected fines. | No. of institutions that denied the information. | Total number of institutions |
| --- | --- | --- | --- |
| 31 [39.3%] | 14 [17.7%] | 34 [43%] | 79 |

With respect to providing information on the presence of vendors within 100 yards of the educational institutions, 24 out of the 79 stated that there were no vendors within 100 yards of the educational institution. 3 out of the 79 stated that there was a presence of vendors selling cigarettes and tobacco products within 100 yards of the educational institution. 52 educational institutions denied the information or stated that there was no such record (with others stating that it was beyond their purview to have a record). See Table 6.

**Table 6:**
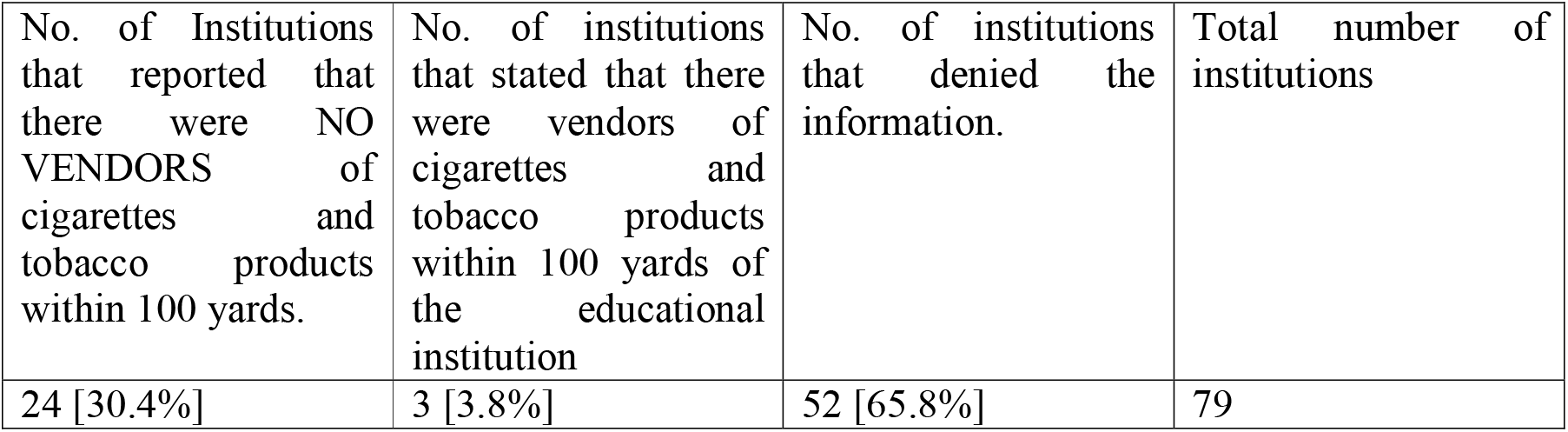
The summary of the responses by institutions about the number of vendors of cigarettes and other tobacco products within a distance of 100 yards of the educational institute.

The information pertaining to other steps taken by institutions to prevent the use of cigarettes and tobacco products by students in the educational institutions was asked for. 57 of the 79 institutions stated that they performed some kind of awareness activity. This started from the bare minimum of issuing a circular to some creating committees and other performing activities, celebrating No Tobacco Day etc. Some went all the way in sending complaints to local law enforcement authorities in case they saw the presence of vendors in their proximities. Five institutions out of the total provided no information on any activity performed, which may mean that no activity has been performed related to awareness of smoking and tobacco use among students of the institution. 17 out of the 79 did not provide the information.

**Table 7:** Summary of the number of institutions that provided a list of events/initiatives/activities/circulars/anything on record where the educational institution has taken steps to prevent the use of cigarettes and tobacco products.

| No. of Institutions that stated they performed actions related to the prevention of smoking on campus. | No. of institutions that stated that provided no information on any activity performed. This may mean no activity was performed. | No. of institutions that denied the information. | Total number of institutions |
| --- | --- | --- | --- |
| 57 [72.2%] | 5 [6.3%] | 17 [21.5%] | 79 |

## Discussion

Many studies related to this present study, but focussing on the presence of vendors of tobacco/cigarettes within 100 yards of educational institutions, have been performed in India [9–12]. This includes studies in Mohali (Punjab, Vadodara (Gujarat) and Chennai (Tamil Nadu) [13]. There was one that was performed in Delhi but had a sample size of only 10 per cent of the schools in Delhi, and the schools were selected randomly[14]. Another Indian study was performed but was limited to the surroundings of hospital buildings as institutions, which reported a high presence of tobacco vendors [15]. But this present study is all the more important and relevant for a list of reasons. This includes the large number of sample data, which is 79 educational institutions. Another important factor is that the educational institutions in this study are scattered throughout the territory of India, and this prevents any kind of regional limitation in this study. A pertinent factor making this study unique is that it has been performed using India’s transparency lay, i.e., the Right to Information Act, 2005[16], which ensures that the information is most reliable. This has been used in previous studies from other government institutions in India [17–19]. This is because the information has been provided by the educational institutions themselves, that too by a senior ranking officer of the institution responsible for the actual implementation, and is provided under seal and signature of the educational institution. This nullifies the presence of any error or bias, and the data is of high integrity. On the possibility of the educational institutions providing incorrect information, it is less probable as the information has been provided in writing under law, and the officer in the institution is deemed to be aware of the consequences, which include fine and disciplinary action under the Right to Information Act, 2005 and the Indian Penal Code.

In the study, it was found that only 31 per cent of the educational institutes had put the boards in the proper format, which prohibited the sale of cigarettes and tobacco products within 100 yards of the educational institutes. As far as the No Smoking boards were concerned, here too, only 58% or 6 in 10 “institutes of national importance” had taken the simple action of placing boards on their campus.

The effect of boards on people has a positive effect as it is an anti-tobacco health message [20]. But regardless of the effect, it is part of the legislation and forms part of the law of the land in India, and non-compliance is a violation.

The ill effects of smoking and tobacco have been reiterated by studies[21], but the relevance of this particular study is to the use of smoking and tobacco by young adults as it relates to higher educational institutions. The use of cigarettes and tobacco at an early age affects lung health at a later age and may also serve as an impediment to quitting at an early age [22].

It has shown in studies that the initiation of tobacco in India is at an early age [23]. This justifies the actions specified in the Indian Anti-smoking law and its related rules, which provide for actions in educational institutes where young people are present. This is all the more necessary as the vendor, with commercial interest, usually does not ask the age of the people they are selling tobacco products [24]. The prevalence of tobacco sellers within 100 yards, which may be non-permanent, ‘shack’ based, is prevalent [25,26].

With respect to the implementation of fines by educational institutions, the paper brings to light the concept where the implementation of the smoking law has been decentralised as the authority to collect fines and compound the offence has been distributed from law enforcement agencies to the people who are running the individual buildings or other public places. The Schedule III of the Prohibition of Smoking in Public Places Rules, 2008[6] gives the power to the head of educational institutes, hospital superintendents in hospitals, postmasters in post offices, station in charge in bus stands, librarians in libraries, etc. The intent is to decentralise and make it a grassroots movement. The non-compliance by India’s leading educational institutes shows a gap in the implementation of the law in India.

With respect to compliance with fines, 14 per cent of the institutions actually had recorded incidences and collected fines or simply issued warnings. Another 31 simply stated that no instances were reported, and hence no fines were imposed. In the Indian condition, more studies are required, and more compliance is required to actually check whether there were actually no incidences on the campus as there is considerable cigarette and tobacco use among youth in India [27]. People may argue that the anti-smoking law in India may have less ‘teeth’ and may not be very stringent, but with respect to the sale of tobacco products to minors, even the Indian legislature has taken the lead and made such act a serious punishment with jail time upto seven years under the Juvenile Legislation in India [7]

But there is one area where the educational institutes in this study have actually taken action. 57% of the institutes had actually taken some form of prevention action or the other. This may include issuing circulars, creating committees, holding counselling sessions, performing skits on tobacco-related topics, celebrating No Tobacco Day, and undertaking inspections.

It is also important to highlight another critical factor in this study, and that is the use of India’s transparency law to gather the information. Around ten institutes did not comply with the information requests fully under the Right to Information Act 2005. This means that compliance with the law is under question and an issue of concern.

Overall this study is a wake-up call for the academic institutions in India that are the ultimate torchbearers of the intention of the legislature to turn the anti-smoking and tobacco law into a reality.

## Conclusion

The study set out to find the compliance status of the anti-smoking/tobacco law in India’s premier higher educational institutions. From the study of 79 institutes of national importance that have been covered in this study, It has been shown with some confidence that compliance with India’s anti-smoking/anti-tobacco law is not universal and severely lacks on many grounds. These include non-compliance with something as basic as the installation of boards within and outside of the campus prohibiting the sale/use of cigarettes and tobacco products. The five objects of the study have been tested, and in all areas, there is serious concern regarding the full implementation of the COTPA Act 2003. As a decentralised law, it is up to these educational institutes, which form an ‘elite group’ of publically funded higher class institutes and christen themselves as institutes of national importance in India.

## Data Availability

Data available on reasonable request

## Declarations

The author declares no conflict of interest/competing interests. No specific funding was taken for this study.

## Acknowledgements

Special thanks to all the Public Information Officers across educational institutions covered in the study, that provided the information in a timely manner. Special thanks to Tathatara Foundation and ISAC Centre for Built Environment Policy for their support. Special thanks to Sandeep K.

## Footnotes

1 ‘beedi’ is an Indian country cigarettes made with tobacco placed inside a rolled dried leaf and in cheaper in price compared to a cigarette.

## References

1 Institutions of National Importance. https://www.education.gov.in/en/institutions-national-importance (accessed 14 Feb 2023).

2 Institutes of National Importance: Wikipedia. https://en.wikipedia.org/wiki/Institutes_of_National_Importance (accessed 14 Feb 2023).

3 World Health Organisation. Tobacco or Health: Agenda Item 22; Resolutions of the Thirty-Ninth World Health Assembly of Interest to the Regional Committee. https://iris.paho.org/bitstream/handle/10665.2/30360/22_10.pdf?sequence=1&isAllowed=y. (accessed 14 Feb 2023).

4 Government of India. The Cigarettes and other Tobacco Products (Prohibition of Advertisement and Regulation of Trade and Commerce, Production, Supply and Distribution) Act, 2003. 2003. https://legislative.gov.in/sites/default/files/A2003-34.pdf (accessed 14 Feb 2023).

5 Ministry of Health and Family Welfare, Government of India. Cigarettes and other Tobacco Products (Display of Boards by Educational Institutions) Rules, 2009. https://nhm.gov.in/cota/Notifications%20on%20Section-6%28b%29 x(accessed 14 Feb 2023).

6 Government of India. The Prohibition of Smoking in Public Place Rules, 2008. 2008. https://nhm.gov.in/cota/Notifications%20on%20Section%204%20of%20the%20Act%20Related%20to%20Prohibition%20of%20Smoking%20in%20Public%20Places/GSR-417-D%28E%29.pdf (accessed 14 Feb 2023).

7 Government of India. The Juvenile Justice (Care and Protection of Children) Act, 2015. 2016. https://legislative.gov.in/sites/default/files/A2016-2_0.pdf (accessed 14 Feb 2023).

8 National Ethics Guidelines for Biomedical and Health Research involving Human Participants. Indian Council of Medical Research 2017. https://ethics.ncdirindia.org//asset/pdf/ICMR_National_Ethical_Guidelines.pdf (accessed 6 Mar 2022).

9 Mistry R, Pednekar M, Pimple S, et al. Banning tobacco sales and advertisements near educational institutions may reduce students’ tobacco use risk: evidence from Mumbai, India. Tobacco Control 2015;24:e100–7. doi:10.1136/tobaccocontrol-2012-050819

10 Khargekar NC, Debnath A, Khargekar NR, et al. Compliance of Cigarettes and Other Tobacco Products Act among Tobacco Vendors, Educational Institutions, and Public Places in Bengaluru City. Indian Journal of Medical and Paediatric Oncology 2018;39:463–6. doi:10.4103/ijmpo.ijmpo_136_17

11 Mistry R, Pednekar MS, McCarthy WJ, et al. Compliance with point-of-sale tobacco control policies and student tobacco use in Mumbai, India. Tob Control 2019;28:220–6. doi:10.1136/tobaccocontrol-2018-054290

12 Elf JL, Modi B, Stillman F, et al. Tobacco sales and marketing within 100 yards of schools in Ahmedabad City, India. Public Health 2013;127:442–8. doi:10.1016/j.puhe.2013.02.003

13 Goel S, Kumar R, Lal P, et al. How Compliant are Tobacco Vendors to India’s Tobacco Control Legislation on Ban of Advertisments at Point of Sale? A Three Jurisdictions Review. Asian Pacific Journal of Cancer Prevention 2015;15:10637–42. doi:10.7314/APJCP.2014.15.24.10637

14 Bassi S, Gupta VK, Park M, et al. School policies, built environment and practices for non-communicable disease (NCD) prevention and control in schools of Delhi, India. PLoS ONE 2019;14:e0215365. doi:10.1371/journal.pone.0215365

15 Rijhwani K, Mohanty VR, Balappanavar AY, et al. Compliance Assessment of Cigarette and Other Tobacco Products Act in Public Places in Delhi Government Hospitals. Asian Pac J Cancer Prev 2018;19. doi:10.22034/APJCP.2018.19.8.2097

16 Singh R. RTI for Research: Using the Right to Information Act, 2005 for Research in India. New Delhi: : Sandeep Kaur(BooksBonanza) 2020. 10.5281/zenodo.6088938

17 Singh, Raja, Dewan, Anil. Air conditioners, airborne infection prevention and air pollution in buildings in New Delhi. International Journal of Tuberculosis and Lung Diseases 2022;26:288–90. doi:http://dx.doi.org/10.5588/ijtld.21.0704

18 Singh R. Studying the Double Paradox in Air Conditioning at Indian Airports for Airborne Infection Prevention and Filtration of Harmful Suspended Particulate Matter. Cureus 2022;14. doi:10.7759/cureus.23748

19 Singh R. The Risk Status of Waiting Areas for Airborne Infection Control in Delhi Hospitals. Cureus 2022;14.

20 Laxmi Kumari, Meenakshi Sood, Sandhya Gupta. Effect of anti-tobacco health messages post-implemetation of COTPA 2003 in India. Indian J Psy Nsg 2022;19:163–70. doi:10.4103/iopn.iopn_66_22

21 Tager IB, Muñoz A, Rosner B, et al. Effect of cigarette smoking on the pulmonary function of children and adolescents. Am Rev Respir Dis 1985;131:752–9. doi:10.1164/arrd.1985.131.5.752

22 Khuder SA, Dayal HH, Mutgi AB. Age at smoking onset and its effect on smoking cessation. Addictive Behaviors 1999;24:673–7. doi:10.1016/S0306-4603(98)00113-0

23 Singh V, Pal HR, Mehta M, et al. Tobacco consumption and awareness of their health hazards amongst lower income group school children in National Capital Territory of Delhi. Indian Pediatr 2007;44:293–5.

24 Verma AR. Knowledge, Attitude, and Practices of Tobacco Vendors toward Selling Tobacco Products to Young Children and Adolescents in Central Delhi. International Journal of Clinical Pediatric Dentistry 2021;14:97–9. doi:10.5005/jp-journals-10005-1914

25 Kumar S, Kapoor S, Sharma R, et al. Compliance assessment with tobacco control regulations at wheelchair-based tobacco Point of sale in Delhi, India. Int J Non-Commun Dis 2021;6:38. doi:10.4103/jncd.jncd_76_20

26 Singh DrR. Signboards Prohibiting Tobacco Sale Within 100 Yards of Educational Institutes: The Appraisal of Prohibition Compliance and On-Ground Status of the Anti-smoking Law in New Delhi’s Major Administrative Precinct. 2023. doi:10.32388/KU2Z0X.3

27 Grover S, Anand T, Kishore J, et al. Tobacco Use Among the Youth in India: Evidence From Global Adult Tobacco Survey-2 (2016-2017). Tob Use Insights 2020;13:1179173X2092739. doi:10.1177/1179173X20927397

